# Vaccine acceptance among college students in South Carolina: Do information sources and trust in information make a difference?

**DOI:** 10.1101/2020.12.02.20242982

**Authors:** Shan Qiao, Daniela B. Friedman, Cheuk Chi Tam, Chengbo Zeng, Xiaoming Li

## Abstract

**Background:** To control the COVID-19 pandemic, governments need to ensure a successful large-scale administration of COVID-19 vaccines when safe and efficacious vaccines become available. Vaccine acceptance could be a critical factor influencing vaccine uptake. Health information has been associated with vaccine acceptance. For college students who are embracing a digital era and being exposed to multimedia, the sources of COVID-19 vaccine information and their trust in these sources may play an important role in shaping their acceptance of vaccine uptake.

**Methods:** In September 2020, we conducted an online survey among 1062 college students in South Carolina to understand their perceptions and attitudes toward COVID-19 vaccination. Descriptive analysis and linear regression analysis were used to investigate vaccine information sources among college students and examine how COVID-19 vaccine acceptance was associated with information source and trust level in each source.

**Results:** The top three sources of COVID-19 vaccine information were health agencies (57.7%), mass media (49.5%), and personal social networks (40.5%). About 83.1% of the participants largely or always trusted scientists, 73.9% trusted healthcare providers, and 70.2% trusted health agencies. After controlling for key demographics, vaccine acceptance was positively associated with scientists as information sources but negatively associated with pharmaceutical companies as sources. Higher trust levels in mass media, health agencies, scientists, and pharmaceutical companies was significantly associated with higher COVID-19 vaccine acceptance. However, trust in social media was negatively associated with vaccine acceptance.

**Discussion:** College students use multiple sources to learn about upcoming COVID-19 vaccines including health agencies, personal networks, and social media. The level of trust in these information sources play a critical role in predicting vaccine acceptance. Trust in health authorities and scientists rather than social media is related to higher level vaccine acceptance. Our findings echo the call for restoring trust in government, healthcare system, scientists, and pharmaceutical industries in the COVID-19 era and highlight the urgency to dispel misinformation in social media. Effective strategies are needed to disseminate accurate information about COVID-19 vaccine from health authorities and scientific research to improve vaccine communication to the public and promote COVID-19 vaccine uptake.

## Introduction

The COVID-19 pandemic has severely disrupted normal societal and economic activities worldwide and is expected to continue imposing strains and burden on health systems in most of countries. Encouraging news is that several vaccines are in phase 3 clinical trials and showing promising effectiveness ^1^. When safe and efficacious vaccines become available, policy makers need to ensure a successful large-scale uptake of COVID-19 vaccines to achieve community immunization. Vaccine hesitancy, referred as “a delay in acceptance or refusal of vaccination despite availability of vaccination services” ^2^, has been recognized as one of the top ten global health threats in 2019 ^3^. Existing studies regarding COVID-19 vaccine uptake suggest that a considerable proportion of people are reluctant to get vaccination against COVID-19. A global survey of COVID-19 vaccine acceptance in 19 countries reported that 71.5% of the participants would be very or somewhat likely to take a COVID-19 vaccine ^4^. A systematic review on acceptance of a COVID-19 vaccine based on nationally representative surveys in 20 nations suggests that the vaccine acceptance rate in most of the nations would not reach the 67% necessary for achieving population immunity ^5^.

Extant literature demonstrates that the determinants of vaccine acceptance are multiple, complex, and vary depending on type of vaccine is involved ^6^. The lessons learned from previous infectious disease outbreaks, including SARS, H1N1, and Ebola demonstrate the important role that health information has on disease control and vaccine acceptance ^7^. Source of health information can affect the manner and frequency of the utilization of such information. The degree to which the information source is trusted can have a markable impact on the acceptance of information ^8^. If people distrust the source, they will doubt the information, and this doubt will in turn shape their attitudes, perceptions, and potential actions they take (such as vaccine uptake) ^9-11^.

A growing literature focuses on sources and perceived credibility of vaccine information as well as their impacts on vaccine acceptance. One study on HPV vaccine uptake in Georgia, United States suggested that sources of information about HPV vaccine were associated with parental vaccine acceptance and further influenced vaccine uptake among adolescents ^12^. Another study conducted in France showed that vaccine acceptance was higher when patients reported getting information from healthcare providers than from the Internet or relatives ^13^. Moran and colleagues investigated the association between information sources, trust in sources, and vaccine safety concerns among women of different ethnicities in Los Angeles, reporting that both the information sources and trust in health information resources were associated with vaccine safety concerns, but the patterns of these associations varied by ethnicity ^14^.

Researchers have expressed concerns regarding the infodemic that has emerged during the COVID-19 outbreak ^15, 16^ and warned that misinformation and conspiratorial beliefs spreading through various channels may considerably reduce COVID-19 vaccine acceptance ^17^. There is a dearth of studies that investigate the sources of information about COVID-19 vaccines, assess how people trust vaccine information from different sources, or explore how the sources of information and trust in these sources affect their acceptance of COVID-19 vaccines. In addition, most studies on vaccine acceptance and health information are conducted with parents since the vaccines involved are related to childhood vaccination and parents play a dominant role in decision making regarding their children’s vaccine uptake. In the context of COVID-19, it is important to engage young adults such as college students in the vaccine campaigns and understand where they receive vaccine-related information, their trusted sources of COVID-19 vaccine, and how these information sources and trust in vaccine information from different sources shape their acceptance of vaccine uptakes. College students are susceptible to health-compromising behaviors due to a sense of invulnerability ^18^, comparative optimism, and a perception that COVID-19 is not a serious health threat ^19^. Being part of a generation that embraces a digital era and is exposed to multimedia, college students may be more influenced by online information and social media in their decision making about getting vaccinated against COVID-19.

The current study aims to explore the information sources of COVID-19 vaccine among college students, assess their trust in different information sources, and examine how the sources and trust of COVID-19 vaccine information are associated with the acceptance of a COVID-19 vaccine after controlling for key demographics.

## Methods

### Participants and procedure

An online survey was conducted between September and October 2020 among college students in South Carolina. The participants were recruited through a convenience sampling approach with inclusion criteria including: (1) being 18 years of age or older; and (2) being currently a full-time student enrolled in a university. Specifically, an email invitation was distributed to student listservs by various colleges (e.g., School of Public Health, School of Nursing, School of Social Work, etc.) and departments on campus. The invitation email included a weblink of the survey and an online consent covering study purposes, procedure, voluntary nature, and confidentiality protection. The survey was developed using RedCap ^20^, a widely used online platform in health surveys. The self-administered and anonymous survey typically took about 20 minutes to complete. The participants were also encouraged to share the invitation and the survey link with other students. All participants were provided with an option to enter a prize drawing to win a $25 Amazon e-gift card. Ten e-gift cards were given away through a random drawing. A total of 1,370 college students participated in the survey. Data from 308 participants were removed due to incomplete responses (i.e., finishing less than half of the survey). The final sample size of the current study was 1,062. The research protocol was approved by the Institutional Review Board at the University of South Carolina.

### Measures

#### Sociodemographic characteristics

Participants were asked to provide their sociodemographic characteristics including gender, age, race/ethnicity, college year, and annual family income. Given that certain categories included very few participants (< 5%), two variables were dichotomized, including gender (0 = Female, 1 = Male) and race/ethnicity (0 = non-Caucasian, 1 = Caucasian) for data analyses.

#### Vaccine acceptance

One question was used to assess participants’ likelihood to get a COVID-19 vaccine (i.e., “How likely will you get a COVID-19 vaccine when it is available”). Participants responded to this question on a five-point Likert scale (1 = definitely not take it, 2= not likely to take it, 3 = I don’t know, 4 = likely to take it, and 5 = definitely take it). In the descriptive data analysis, we further categorized participants into three groups based on their responses: (1) refusal group (participants with answers of ‘1’ or ‘2’); (2) hesitancy group (participants with answers of ‘3’); and (3) acceptance group (participants with answers of ‘4’ or ‘5’).

#### Sources of COVID-19 vaccine information

Participants were asked where they would typically obtain information about the COVID-19 vaccine. The response items were: 1) I do not get any information from anywhere; 2) social media; 3) mass media (TV, newspapers); 4) government; 5) health agencies; 6) scientists; 7) pharmaceutical companies (e.g., vaccine producers); 8) my healthcare providers; 9) my personal social network (friends, classmates, or teachers); and 10) other source. The response option was “yes” or “no” for each item.

#### Trust in information sources about the COVID-19 vaccine

Participants were asked about their levels of trust in COVID-19 vaccine information from each of the following sources: social media, mass media, government, health agencies, scientists, pharmaceutical companies, their healthcare providers, and their personal social network. For each information source, participants could respond on a five-point scale ranging from “Not trust at all” (1) to “Always trust” (5). In the descriptive analysis, this variable was dichotomized into “not trust at all/very little trust/average trust” and “largely trust/always trust”.

### Data analysis

Descriptive statistics were reported on sociodemographic variables, COVID-19 vaccine acceptance, sources of vaccine information, and trust in sources by vaccine acceptance groups (refusal, hesitancy, and acceptance). ANOVA for continuous variables and chi-square tests for categorical variables were used to examine any differences in demographics, information sources, and trust in information sources by COVID-19 vaccine acceptance groups (i.e., refusal, hesitancy, and acceptance).

Treating vaccine acceptance as a continuous variable, we conducted linear regression modeling on vaccine acceptance score to examine the association of information sources, trust in information sources with COVID-19 vaccine acceptance, controlling for sociodemographic characteristics. Standardized regression coefficient (*β)* were reported for each predictor in the regression model. All statistical analyses were performed using SPSS software version 26 ^21^.

## Results

### Descriptive statistics and univariate analysis

As shown in Table 1, participants were 23.83 years of age on average (*standard deviation* [*SD*] = 6.66). Most participants were female (79.8%), Caucasian (85.9%), with annual family income of $50,000 to $100,000. More than half of the participants were undergraduates (17.1% Senior, 12.4% Junior, 12.2% Freshmen, and 10.5% Sophomore) with 27.6% being doctoral students and 19.5% master students.

**Table 1.** Demographic analyses between COVID-19 vaccination acceptance groups (*n* = 1062)

|  | Overall | Refusal group | Hesitancy group | Acceptance group | Group comparisons |  |
| --- | --- | --- | --- | --- | --- | --- |
|  |  |  |  |  | *F/Chi-square* | *p-value* |
| *n* | 1062 | 258 (24.3%) | 160 (15.1%) | 644 (60.6%) |  |  |
| Age, *Mean (SD)* |  | 24.64 (7.53) | 23.21 (6.03) | 23.64 (6.41) | 2.77 | 0.063 |
| Gender |  |  |  |  |  |  |
| Female | 848 (79.8%) | 214 (83.3%) | 134 (83.8%) | 500 (77.9%) | 4.93 | 0.085 |
| Male | 211 (19.9%) | 43 (16.7%) | 26 (16.3%) | 142 (22.1%) |  |  |
| Race/Ethnicity<sup>a</sup> |  |  |  |  |  |  |
| White/Caucasian | 912 (85.9%) | 216 (83.7%) | 136 (85.0%) | 560 (87.0%) | 1.71 | 0.425 |
| Black/Africa American | 71 (6.7%) |  |  |  |  |  |
| Hispanic/Latino | 32 (3.0%) |  |  |  |  |  |
| Asian | 85 (8.0%) |  |  |  |  |  |
| American Indian/Alaskan Native | 6 (0.6%) | 42 (16.3%) | 24 (15.0%) | 84 (13.0%) |  |  |
| Native Hawaiian/ other Pacific Islander | 3 (0.3%) |  |  |  |  |  |
| Other | 6 (0.6%) |  |  |  |  |  |
| Annual family income |  |  |  |  |  |  |
| < \$10,000 | 63 (5.9%) | 15 (5.8%) | 8 (5.1%) | 40 (6.3%) | 13.23 | 0.104 |
| \$10,000 to \$24,999 | 102 (9.6%) | 15 (5.8%) | 18 (11.5%) | 69 (10.8%) | | |
| \$25,000 to \$49,999 | 161 (15.2%) | 47 (18.3%) | 29 (18.6%) | 85 (13.4%) | | |
| \$50,000 to \$100,000 | 306 (28.8%) | 84 (32.7%) | 46 (29.5%) | 176 (27.7%) | | |
| >\$100,000 | 417 (39.3%) | 96 (37.4%) | 55 (35.3%) | 266 (41.8%) | | |
| School year<sup>b</sup> |  |  |  |  |  |  |
| Freshman | 130 (12.2%) |  |  |  | 3.51 | 0.173 |
| Sophomore | 112 (10.5%) | 143 (55.6%) | 90 (57.0%) | 323 (50.3%) |  |  |
| Junior | 132 (12.4%) |  |  |  |  |  |
| Senior | 182 (17.1%) |  |  |  |  |  |
| Masters student, first year | 207 (19.5%) | 114 (44.4%) | 68 (43.0%) | 319 (49.7%) |  |  |
| Doctoral student, first year | 294 (27.6%) |  |  |  |  |  |
| Types of information sources |  |  |  |  |  |  |
| Social Media | 404 (38.0%) | 91 (35.3%) | 68 (42.5%) | 245 (38.0%) | 2.19 | .335 |
| Mass media | 526 (49.5%) | 115 (44.6%) | 76 (47.5%) | 335 (52.0%) | 4.40 | .111 |
| Government | 205 (19.3%) | 59 (22.9%) | 26 (16.3%) | 120 (18.6%) | 3.24 | .197 |
| Health agencies | 613 (57.7%) | 158 (61.2%) | 86 (53.8%) | 369 (57.3%) | 2.39 | .303 |
| Scientists | 349 (32.9%) | 79 (30.6%) | 44 (27.5%) | 226 (35.1%) | 4.13 | .127 |
| Pharmaceutical companies | 187 (17.6%) | 54 (20.9%) | 23 (14.4%) | 110 (17.1%) | 3.24 | .198 |
| Healthcare providers | 206 (19.4%) | 59 (22.9%) | 28 (17.5%) | 119 (18.5%) | 2.70 | .259 |
| My personal social networks | 430 (40.5%) | 93 (36.0%) | 72 (45.0%) | 265 (41.1%) | 3.58 | .167 |
| Trust of information sources<sup>c</sup> |  |  |  |  |  |  |
| Social Media | 18 (1.7%) | 5 (1.9%) | 1 (0.6%) | 12 (1.9%) | 1.28 | .526 |
| Mass media | 72 (6.8%) | 9 (3.5%) | 8 (5.0%) | 55 (8.5%) | **8.34\*** | .015 |
| Government | 146 (13.8%) | 25 (9.7%) | 19 (11.9%) | 102 (15.9%) | **6.43\*** | .040 |
| Health agencies | 745 (70.2%) | 136 (52.7%) | 104 (65.4%) | 505 (78.4%) | **60.26\*\*\*** | .000 |
| Scientists | 882 (83.1%) | 186 (72.1%) | 121 (76.1%) | 575 (89.3%) | **45.41\*\*\*** | .000 |
| Pharmaceutical companies | 313 (29.6%) | 53 (20.5%) | 40 (25.2%) | 220 (34.3%) | **18.53\*\*\*** | .000 |
| Healthcare providers | 782 (73.9%) | 153 (59.3%) | 105 (66.0%) | 524 (81.7%) | **53.08\*\*\*** | .000 |
| My personal social networks | 133 (12.6%) | 36 (14.1%) | 12 (7.5%) | 85 (13.2%) | 4.42 | .110 |

<sup>a</sup> Race/Ethnicity was dichotomized into 0 (White/Caucasian) and 1 (non-White/Caucasian) for analyses
<sup>b</sup> School year was dichotomized into 0 (undergraduate) and 1 (graduate) for analyses.
<sup>c</sup> Trust of information sources were dichotomized into 0 (not trust at all/very little trust/average) and 1 (largely trust/always trust).
Descriptive statistics were reported among participants who reported to trust in corresponding information sources.
\* $p < .05$ ; \*\*\* $p < .001$

About 60.6% of participants reported that they would likely or definitely to get vaccinated against COVID-19 (acceptance group), 24.3% expressed that they would likely or definitely not take the vaccines (refusal group), and 15.1% had no clear idea about vaccine uptake (hesitancy group). The top three information sources about the vaccine were health agencies (57.7%), mass media (49.5%), and personal social networks (40.5%), followed by social media (38%), scientists (32.9%), healthcare providers (19.4%), government (19.3%), and pharmaceutical companies (17.6%). In terms of trust level for each information source, 83.1% of the participants largely or always trusted scientists, 73.9% trusted healthcare providers, and 70.2% trusted health agencies. About 29.6%, 13.8%, 12.6% of the participants highly trusted pharmaceutical companies, government, and personal social networks, respectively. The participants showed low levels of trust in mass media (6.8%) and social media (1.7%).

Univariate analysis did not detect any statistically significant difference across three COVID-19 vaccine acceptance groups in terms of demographics and information sources about COVID-19 vaccines. However, univariate analysis suggested that the acceptance group demonstrated a significantly higher level of trust in health agencies, healthcare providers, scientists, pharmaceutical companies (*p* <.0001 for all) as well as mass media (*p* = .015) and government (p=.04).

### Multivariate regression

Results of the linear regression model are presented in Table 2. Being male (*β* = 0.07, *p* = 0.022), being younger (*β* = −0.09, *p* = 0.018), and being a graduate student (*β* = 0.10, *p* = 0.013) were associated with higher levels of COVID-19 vaccine acceptance. Controlling for demographics, reporting information source as scientists was significantly associated with higher vaccine acceptance (*β* = 0.08 *p* = 0.024), while learning about COVID-19 vaccines from pharmaceutical companies was associated with lower acceptance of vaccine uptake (*β* = −0.07, *p* = 0.042). In addition, higher level of trust in mass media (*β* = 0.17, *p* < 0.001), health agencies (*β* = 0.12, *p* = 0.004), scientists (*β* = 0.09, *p* = 0.033), and pharmaceutical companies (*β* = 0.11, *p* = 0.002) was significantly related to higher vaccine acceptance while trust in social media was negatively related to vaccine acceptance (*β* = −0.08, *p* = 0.037).

**Table 2.** Regression on COVID-19 vaccine acceptance with types and trust of information sources among college students

| Predictor Variable | *B* | *SE* | $\beta$ | *t* | *p* |
| --- | --- | --- | --- | --- | --- |
| Gender | 0.23 | 0.10 | **0.07\*** | 2.30 | 0.022 |
| Age | -0.02 | 0.01 | **-0.09\*** | -2.36 | 0.018 |
| White/Caucasian | 0.01 | 0.12 | 0.00 | 0.10 | 0.919 |
| Annua family income | 0.02 | 0.03 | 0.02 | 0.54 | 0.588 |
| Graduates | 0.26 | 0.10 | **0.10\*** | 2.50 | 0.013 |
| Types of information sources |  |  |  |  |  |
| Social Media | 0.02 | 0.09 | 0.01 | 0.18 | 0.860 |
| Mass media | 0.05 | 0.09 | 0.02 | 0.56 | 0.577 |
| Government | -0.21 | 0.11 | -0.06 | -1.89 | 0.059 |
| Health agencies | -0.05 | 0.09 | -0.02 | -0.52 | 0.602 |
| Scientists | 0.21 | 0.09 | **0.08\*** | 2.26 | 0.024 |
| Pharmaceutical companies | -0.23 | 0.11 | **-0.07\*** | -2.03 | 0.042 |
| Healthcare providers | -0.16 | 0.10 | -0.05 | -1.54 | 0.123 |
| My personal social networks | 0.14 | 0.09 | 0.05 | 1.62 | 0.106 |
| Trust of information sources |  |  |  |  |  |
| Social Media | -0.14 | 0.07 | **-0.08\*** | -2.09 | 0.037 |
| Mass media | 0.25 | 0.06 | **0.17\*\*\*** | 4.31 | 0.000 |
| Government | 0.03 | 0.05 | 0.02 | 0.62 | 0.534 |
| Health agencies | 0.20 | 0.07 | **0.12\*\*** | 2.92 | 0.004 |
| Scientists | 0.17 | 0.08 | **0.09\*** | 2.13 | 0.033 |
| Pharmaceutical companies | 0.15 | 0.05 | **0.11\*\*** | 3.10 | 0.002 |
| Healthcare providers | 0.12 | 0.07 | 0.07 | 1.80 | 0.072 |
| My personal social networks | -0.06 | 0.05 | -0.04 | -1.10 | 0.272 |

*SE* = Standardized Error. $F=9.09$ $R^2=0.145$ . Source was dichotomous response to a checklist. Trust was continuous as a higher score indicating greater trust in an information source.
\* $p < .05$ ; \*\* $p < .01$ ; \*\*\* $p < .001$

## Discussion

Although there is increasing literature on COVID-19 vaccine acceptance and factors that are associated with people’s willingness to get vaccinated when the vaccine is available, the current study is one of first efforts to investigate information sources of COVID-19 vaccines, trust in different information sources, and the impact of information sources and trust level on vaccine acceptance among college students in South Carolina. Our findings show that college students have multiple information sources for learning about COVID-19 vaccines (e.g., health agencies to personal networks to social media). However, trust level in these information sources plays a critical role in predicting vaccine acceptance. Trust in health authority and scientists is related to higher vaccine acceptance, while trust in social media is negatively associated with vaccine acceptance.

The current study suggests that college students in South Carolina mainly obtained vaccine information from health agencies. This finding is aligned with existing studies that reported health authorities as dominant information source for parents regarding childhood vaccination ^22^. However, other authorities and key stakeholders of vaccine development and distribution including scientists, healthcare providers, and government were not main information sources of COVID-19 vaccine. This pattern may indicate limited availability or widespread dissemination of vaccine-related scientific messaging by healthcare providers and government. In addition, mass media and personal social networks rather than social media were among the top information sources of COVID-19 vaccine, suggesting that we should not discount the role of this more traditional communication among young adults even in the digital era.

It is notable that the rank of information sources about COVID-19 vaccines is not fully accordance with trust level in these information sources. For example, the majority of participants expressed a high level of trust in scientists and healthcare providers even they were not listed as main information sources regarding vaccines. Despite obtaining information about COVID-19 vaccines from personal social networks and mass media, participants reviewed the two sources as less trustworthy. This finding is inconsistent with literature regarding trust and information sources, which argues that people trust sources of health information to which they are always exposed ^8, 23^. However, studies on prostate cancer communication among African American men show that it is not uncommon for people use one information source (e.g., newspaper, family members) but trust others (doctors and health educators) ^24^. One potential interpretation is that the acquisition of COVID-19 vaccine information occurs along with other escalating information about COVID-19 from various sources during a relatively short time period. While some participants might actively seek out COVID-19 vaccine information, a considerable amount of information is encountered in a more passive and less deliberate way. College students who were exposed to an information source may not necessarily trust the specific COVID-19 vaccine information disseminated through this source. Future studies are needed to further examine potential factors that influence their judgment such as health literacy, health beliefs, and social norms ^25^.

Another interesting finding is that obtaining COVID-19 vaccine information from pharmaceutical companies was associated with lower acceptance of vaccine uptake. There were numerous unverifiable, unaccountable, and often mixed information from pharmaceutical companies regarding the vaccine in the earlier stages of the pandemic. The accelerated pace of vaccine development may further exaggerate public anxieties, confusion, and doubt ^26^. Pharmaceutical companies might be viewed as a negative source on COVID-19 vaccine and impede people’s vaccine acceptance.

Consistent with other studies on vaccine acceptance, our findings highlight the critical role of trust in decision making regarding vaccination ^27-29^. Biomedical science and research act as foundations of vaccine development; pharmaceutical companies develop and produce the vaccines; and health authorities such as the Food and Drug Administration and the Centers for Disease Control and Prevention regulate vaccine production and promote vaccination. Trust in these key stakeholders in the vaccine development continuum was significantly associated with higher level vaccine acceptance among college students. Trust in the health care system, science and technology, and healthcare professionals could be stronger drivers of vaccine acceptance during public health emergencies such as the COVID-19 pandemic when people have to face partial, inconsistent, conditional, and even contradictory information and knowledge about a new virus ^30^. This trust, however, could be fragile when encroached by misinformation and “antivax” activities through social media and personal networks. For example, our study suggests that trust in social media was negatively related to vaccine acceptance.

The current study is subject to several limitations. First, its cross-sectional design limits our exploration of the complicated causal pathways between information sources, trust, and vaccine acceptance. Second, participants in the current study showed a high level of homogeneity in demographics. Although our data did not suggest a racial/ethnic difference in COVID-19 vaccine acceptance, existing literature demonstrates racial differences in trust relationships with health authorities due to historical events and narratives ^27, 31^. Comparison studies are warranted for further exploring the role of race/ethnicity in relation to trust in various information sources and vaccine acceptance. Third, due to the brevity of online survey, data were not available on some other factors that may either mediate or moderate the association between trust in sources of vaccine information and vaccine acceptance. For example, some studies have indicated a potential association between political orientation and vaccine hesitancy ^32^. Fourth, the sample in the current study was from one college in one southeastern state and findings cannot be generalized to other college populations elsewhere. Although growing literature on trust and vaccine hesitancy suggests some commonalities, the relationship may vary by population, vaccine, and region ^33^.

Despite these limitations, the current study elaborates the pattern of sources regarding COVID-19 vaccine information and trust in these information sources among college students in South Carolina as well as explored the impacts of trust on vaccine acceptance, which can inform specific strategies for COVID-19 vaccine promotion among this group. First, we need to increase the coverage of authorized health communication on COVID-19 vaccine among young adults. Scientists and healthcare providers were both trusted sources for vaccine information for college students, yet they were not their main sources of vaccine information. This discrepancy implies a gap in critical communication and education by trusted sources. Scientists and healthcare professionals should have the opportunity to play an active role in disseminating their research findings and dispelling misinformation and conspiratorial messages about COVID-19 vaccine.

Second, as the developer and producer of vaccines, pharmaceutical companies have the responsibilities to provide clear and accurate communication to reduce confusion and doubt and rebuild a trusting relationship with the public. Tarnished reputations of pharmaceutical companies have fueled the anti-vaccine movement ^34, 35^. Playing a positive role in health communication and partnering with health care agencies and scientists on this communication will help address distrust issues with the public.

Third, we need to build increased trust in government in terms of vaccine communication. According to our results, government was neither a main nor a trusted source of COVID-19 vaccine information. Public hesitancy may be intensified by contradictory information from federal and state governments and politicization of vaccine development and approvals. A transparent and evidence-based policy is required to strengthen the public’s trust in government.

Fourth, we need to pay attention to the role of mass media and personal social networks in vaccine communication. Both were reported as main sources of vaccine information among college students in our study, which implies that we should not discount the role of traditional communication sources among young adults even in this digital era. The participants in our study did not show high trust in mass media and personal social networks. Despite people’s distrust in a particular information source, exposure to its content could still induce emotional reactions and sow confusion and doubt ^36^. Similarly, we also need to effectively track, monitor, and disperse misinformation in social media ^37^.

In summary, our findings echo the call for restoring trust in the healthcare system, scientists, and biomedical industries and highlight the urgency to dispel misinformation about COVID-19 vaccines in social media ^38^. Effective strategies are needed to improve trust in pharmaceutical companies and government regarding vaccine communication and encourage these entities to partner with sources already trusted by the public.

## Data Availability

Please contact the corresponding author for the availability of data referred to in the manuscript.

## Acknowledgment

We greatly appreciate the advice and comments from Drs. Bankole Olatosi, Sharon Weissman, and Helmut Albrecht on survey development. Research reported in this publication was supported by the National Institutes of Health under Award Number of NIH R01MH0112376-3S1 and R01AI127203-4S1. The content is solely the responsibility of the authors and does not necessarily represent the official views of the National Institutes of Health.

